# Nonutility of procalcitonin for diagnosing bacterial pneumonia in COVID-19

**DOI:** 10.1101/2022.03.29.22272960

**Authors:** Avi J. Cohen, Laura R. Glick, Seohyuk Lee, Yukiko Kunitomo, Derek A. Tsang, Sarah Pitafi, Patricia Valda Toro, Ethan Zhang, Rupak Datta, Charles S. Dela Cruz, Samir Gautam

**Author notes:** These authors contributed equally to the manuscript and will list their names first on their respective CVs (Laura R. Glick, Seohyuk Lee, Yukiko Kunitomo). **Funding information:** T32 – National Institute of Allergy and Infectious Diseases (AI007517), Claude D. Pepper Older Americans Independence Center at Yale (P30AG021342) and Yale Center for Clinical Investigation Scholar Award to RD; U.S. Department of Veterans Affairs Merit award (BX004661), U.S. Department of Defense (PR181442), and U19 – National Heart, Lung, and Blood Institute (AI089992-09S2) to CSD. T32 – National Heart, Lung, and Blood Institute (HL007778), F32 – National Heart, Lung, and Blood Institute (HL154641), Cystic Fibrosis Foundation Fellowship GAUTAM20D0, Parker B. Francis Fellowship, Robert E. Leet and Clara Guthrie Patterson Trust Mentored Research Award, and Yale Center for Clinical Investigation Scholar Award to SG. **Disclosures:** Samir Gautam, MD PhD – Advisory Board – AstraZeneca.

## Abstract

Patients hospitalized with COVID-19 are at significant risk for superimposed bacterial pneumonia. However, diagnosing superinfection is challenging due to its clinical resemblance to severe COVID-19. We therefore evaluated whether the immune biomarker, procalcitonin, could facilitate the diagnosis of bacterial superinfection. To do so, we identified 185 patients with severe COVID-19 who underwent lower respiratory culture; 85 had superinfection. Receiver operating characteristic curve analysis showed that procalcitonin at the time of culture was incapable of distinguishing patients with bacterial infection (AUC, 0.52). We conclude that static measurement of procalcitonin does not aid in the diagnosis of superinfection in severe COVID-19.

## Text

Bacterial pneumonia is an important complication in patients hospitalized with COVID-19. Its incidence approaches 45% in those receiving mechanical ventilation,(1) and it is associated with increased 28-day mortality.(2) However, establishing the diagnosis of bacterial superinfection is challenging due to its shared clinical features with severe COVID-19, including fever, hypoxemia, and radiographic infiltrates. A further obstacle to diagnosing bacterial infection is the hesitance to obtain lower respiratory cultures (LRCx) due to the potential risk of SARS-CoV-2 aerosolization. Hence, there is considerable interest in identifying effective noninvasive diagnostics for superinfection in COVID-19.

Procalcitonin (PCT) is an inflammatory biomarker that has been used as an adjunctive serum test for bacterial pneumonia for over 20 years.(3) The role of PCT in identifying superinfection during COVID-19 remains equivocal. May *et al*. demonstrated limited diagnostic utility in an analysis of 24 patients with bacterial pneumonia,(4) while other investigators have argued for its efficacy.(5–7) Notably, these studies were limited to patients with co-infection upon presentation to the hospital, which is rare compared to secondary bacterial pneumonia, defined as occurring > 48 hours after admission.(9) In addition, the reported cohorts included patients with varying disease severity, which may confound the interpretation of PCT in COVID-19.(10) In line with this hypothesis, we have demonstrated that PCT levels are associated with clinical severity in non-COVID respiratory viral infection.(11)

Therefore, we sought to evaluate the diagnostic utility of PCT for bacterial superinfection in COVID-19 – specifically in hospitalized patients with severe disease. To do so, we performed a retrospective cohort study of adult patients admitted to Yale New Haven Hospital between January 3, 2020 and July 1, 2020 with severe COVID-19, as defined by the NIH COVID-19 Treatment Guidelines: positive SARS-CoV-2 nucleic acid amplification testing and hypoxemia (SpO_2_ ≤ 94% and/or use of supplemental oxygen).(12) We subsequently identified the subgroup of 185 patients in whom LRCx was collected; 85 demonstrated bacterial superinfection. If multiple specimens were obtained, the index culture was used for analysis. Of the 185 LRCx, 106 were sputum samples and 79 were tracheal aspirates. Patients with positive fungal LRCx and those with positive blood or urine cultures were excluded to restrict the analysis to bacterial respiratory infection.

Data were collected by the Joint Data Analytic Team and verified via manual chart review. Unless otherwise indicated, lab values used for analysis were those closest to the time of LRCx. If no lab value was recorded within 36 hours of LRCx, it was recorded as missing. Data were missing at a rate of less than 15% for all variables. Statistical analysis was conducted using the R software environment (V.3.6.0, Vienna, Austria). To determine significance (defined as *p* < 0.05), the Chi-squared test or Kruskal-Wallis test was performed due to non-normal data distributions. Spearman’s rank correlation coefficient was utilized for regression analysis due to the likelihood of non-linear relationships between biomarkers. The study was approved by Yale’s institutional review board (approval #2000023067). Informed consent was not required due to the non-interventional study design.

This approach identified 1308 patients hospitalized with COVID-19 during the study period. Of these, 185 had both severe COVID-19 and a LRCx; 100 had no bacterial growth and 85 had culture-proven superinfection. As shown in Table 1, these groups were well-matched in terms of demographic and clinical variables. Importantly, the groups demonstrated similar disease severity as indicated by organ failure, intensive care unit (ICU) admission, and mortality. In addition, >70% of LRCx were obtained after 48 hours of admission, thus focusing the analysis on secondary bacterial pneumonia. The distribution of bacterial etiologies showed a predominance of *Staphylococcus aureus* (34.7%) and *Pseudomonas aeruginosa* (12.2%), consistent with prior reports.(4, 9)

**Table 1.** Demographic, clinical, and laboratory characteristics of COVID-19 patients with bacterial superinfection compared to those with negative lower respiratory culture (LRCx)

|  | Pure viral infection<br>(*n* = 100) | Bacterial superinfection<br>(*n* = 85) | *p*-value |
| --- | --- | --- | --- |
| Age, median (IQR) | 64.50 (55.00, 76.25) | 63.00 (49.00, 69.00) | 0.128 |
| Female sex, *n* (%) | 34 (34.0%) | 23 (27.1%) | 0.308 |
| BMI, median (IQR) | 28.22 (23.92, 35.78) | 30.28 (25.63, 36.14) | 0.353 |
| ICU admission, *n* (%) | 84 (84.0%) | 75 (88.2%) | 0.409 |
| Mechanical ventilation, *n* (%) | 57 (57.0%) | 68 (80.0%) | <0.001 |
| Renal failure, *n* (%) | 67 (67.0%) | 65 (76.5%) | 0.156 |
| Shock, *n* (%) | 73 (73.0%) | 66 (79.5%) | 0.304 |
| Mortality, *n* (%) | 22 (22.0%) | 28 (32.9%) | 0.095 |
| LRCx >48h after admission | 65 (65.0%) | 68 (80.0%) | 0.024 |
| Biomarkers at time of LRCx |  |  |  |
| PCT, median (IQR) | 0.40 (0.13, 0.76) | 0.30 (0.14, 0.88) | >0.999* |
| WBC, median (IQR) | 8.30 (4.80, 13.60) | 10.20 (6.00, 14.10) | >0.999* |
| Creatinine, median (IQR) | 1.23 (0.82, 2.48) | 1.06 (0.71, 1.79) | >0.999* |
| C-reactive protein, median (IQR) | 91.85 (30.08, 153.47) | 49.75 (15.05, 108.25) | 0.289 |
| D-Dimer, median (IQR) | 2.02 (1.13, 5.00) | 2.59 (0.93, 4.61) | >0.999* |
| Ferritin, median (IQR) | 1143.00 (653.50, 1982.25) | 816.00 (332.50, 1689.75) | 0.210 |
| Temp, median (IQR) | 98.20 (97.80, 99.23) | 98.90 (98.00, 99.75) | 0.156 |
| Microbiological data |  | Count |  |
| *Staphylococcus aureus*, *n* (%) |  | 34 (34.7%) |  |
| Methicillin-resistant *S. aureus*, *n* (%) |  | 6 (6.1%) |  |
| *Pseudomonas aeruginosa*, *n* (%) |  | 12 (12.2%) |  |
| *Haemophilus influenzae*, *n* (%) |  | 8 (8.2%) |  |
| *Klebsiella pneumoniae*, *n* (%) |  | 7 (7.1%) |  |
| *Enterobacter* spp, *n* (%) |  | 7 (7.1%) |  |
| *Escherichia coli*, *n* (%) |  | 6 (6.1%) |  |
| Beta-hemolytic *streptococcus*, *n* (%) |  | 5 (5.1%) |  |
| *Moraxella catarrhalis*, *n* (%) |  | 4 (4.1%) |  |
| Other bacterial pathogens<sup>†</sup>, *n* (%) |  | 9 (9.2%) |  |
| Polymicrobial infection, *n* (%) |  | 13 (15.3%) |  |
| * Bonferroni-adjusted *p*-values are reported for biomarkers to adjust for multiple comparisons |  |  |  |
| † Other bacterial pathogens include species observed < 3 times. These include *Streptococcus pneumoniae* (2), *Serratia marcescens* (2), *Stenotrophomonas maltophilia* (2), *Citrobacter koseri* (1), *Morganella morganii* (1), *Proteus mirabilis* (1). *p*-values were calculated via chi-squared test or Kruskal-Wallis test, as appropriate. |  |  |  |
| Abbreviations: body mass index (BMI), intensive care unit (ICU), interquartile range (IQR), procalcitonin (PCT), white blood cell count (WBC) |  |  |  |

PCT was similar in patients with pure viral infection compared to those with bacterial superinfection (Fig. 1A). The area under the receiver operating characteristic curve (AUROC) for identifying bacterial pneumonia was 0.52 (95% CI: 0.43-0.60), indicating that PCT is an ineffective test for superinfection in severe COVID-19 (Fig. 1B). Importantly, we noted that 63.2% of patients had received antibiotics within 24 hours of LRCx. To determine whether antibiotic exposure influenced the diagnostic utility of PCT, we evaluated the AUROC in exposed vs unexposed patients. This analysis revealed similarly poor performance in both subgroups: AUC_exposed_ = 0.49; AUC_unexposed_ = 0.50.

**Figure 1.**
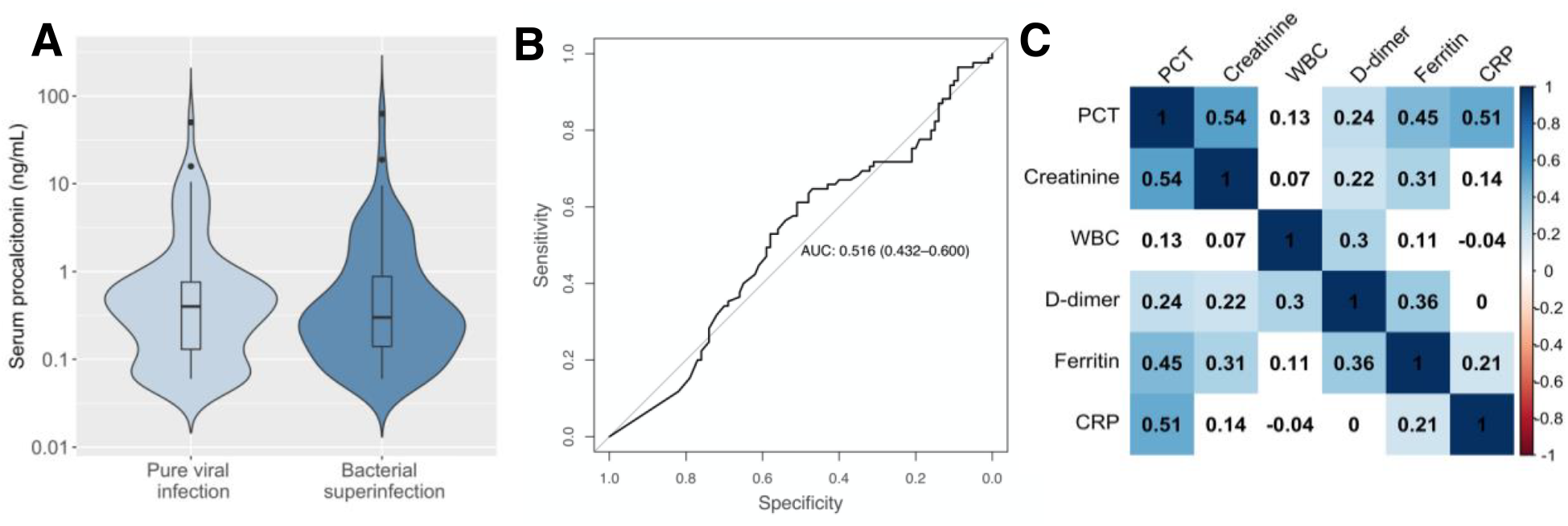
Serum procalcitonin as a biomarker of bacterial superinfection in patients with severe COVID-19. **A**, Procalcitonin levels in patients with pure SARS-CoV-2 infection and those with bacterial superinfection. **B**, Receiver operating characteristic curve defining the utility of PCT in diagnosing superinfection. **C**, Spearman’s rank correlation between biomarkers at the time of LRCx.

Using a cutoff of 0.5 ng/mL, the sensitivity and specificity of PCT were unacceptably low at 33% and 60%, respectively, and the negative predictive value was 51%. PCT, in fact, represented an inferior test for superinfection compared to several non-specific inflammatory biomarkers, including ferritin (Table 2). In contrast, we found that PCT was more effective at predicting severe disease within the study population, as indicated by the AUROC for organ failure, ICU admission, and death.

**Table 2.** Diagnostic performance of procalcitonin and other biomarkers in the identification of bacterial superinfection compared with other clinical outcomes.

| Clinical outcome during admission | Super-infection | Shock | Renal insufficiency | Mechanical ventilation | ICU admission | Mortality |
| --- | --- | --- | --- | --- | --- | --- |
| Biomarker | Area Under the ROC (AUROC) |  |  |  |  |  |
| **PCT** | **0.516** | **0.647** | **0.720** | **0.611** | **0.644** | **0.625** |
| WBC | **0.560** | 0.580 | 0.626 | 0.706 | 0.746 | 0.626 |
| Creatinine | **0.555** | 0.620 | 0.837 | 0.509 | 0.522 | 0.683 |
| C-reactive protein | **0.501** | 0.664 | 0.710 | 0.670 | 0.742 | 0.670 |
| D-Dimer | **0.600** | 0.621 | 0.627 | 0.525 | 0.578 | 0.576 |
| Ferritin | **0.592** | 0.552 | 0.552 | 0.514 | 0.523 | 0.563 |
| Temperature | **0.598** | 0.641 | 0.501 | 0.588 | 0.575 | 0.562 |

Definitions: Shock (Systolic blood pressure < 90); renal insufficiency (creatinine > 1.5)

It is important to note that patients were identified retrospectively in this study according to receipt of LRCx; respiratory cultures were not obtained in all patients. Consequently, these data should not be used to estimate the incidence of bacterial superinfection in our cohort. Additionally, the study was restricted to patients with severe disease, in whom baseline PCT is often elevated.(10) As such, it was not designed to assess the utility of low PCT in ruling out bacterial pneumonia during mild COVID-19.

Taken together, our findings suggest that PCT represents a marker of disease severity rather than bacterial superinfection in COVID-19, as recently shown in influenza and other viral respiratory infections.(11) Accordingly, we propose that PCT be interpreted as a general indicator of the inflammatory host response, which may derive from superinfection or severe COVID-19 itself. Indeed, PCT correlated closely with several other non-specific immune biomarkers in our study including ferritin (Spearman’s ρ = 0.45, *p* < 0.001) and CRP (Spearman’s ρ = 0.51, *p* < 0.001) (Fig. 1C).

In the absence of specific clinical, radiographic, and biochemical tests for bacterial pneumonia in COVID-19, direct microbiologic sampling may represent the optimal means of identifying this important complication.(1) Ongoing evaluation of the safety and efficacy of routine LRCx for the diagnosis of superinfection will be needed.(13)

## Data Availability

All data produced in the present study are available upon reasonable request to the authors.

